# Students’ perspectives on global surgery and its integration into formal medical education

**DOI:** 10.1101/2023.01.03.23284155

**Authors:** Nicholas Rennie, Eva Degraeuwe, Charlotte Deltour, Miryam Serry Senhaji, Margo Vandenheede, Frederik Berrevoet, Elke Van Daele, Wouter Willaert

## Abstract

Global surgery is “an area of study, research, practice, and advocacy that seeks to improve health outcomes and achieve health equity for all people who require surgical care”. The Lancet Commission on Global Surgery strives for an additional 2.28 million physicians in the global surgery workforce by 2030. Understanding how medical students perceive global surgery will be essential in educating the next generation of global surgeons, anaesthesiologists and obstetricians. This study investigated the knowledge, attitudes and exposure of Belgian medical students towards global surgery.

An anonymous online survey was distributed to first to final year medical students across Belgian universities using social media. Data was collected on demographics, exposure, knowledge, obstacles and attitudes of medical students towards global surgery. Odds ratios with 95% confidence intervals were calculated.

A total of 304 medical students participated from four Belgian universities. A minority of students reported having exposure to global surgery (24.7%). Most students were interested in gaining more exposure to global surgery (75.3%). Almost all students agreed (94.4%) that global surgery is a relevant topic for medical students, and most agreed (71%) that there should be more compulsory education on the topic. Only 13 to 44% of students could correctly answer questions testing global surgery knowledge. Students reported multiple barriers to pursuing a career in global surgery. Belgian medical students’ global surgery knowledge and exposure is lacking despite clear interest in the field. Educating and supporting students will be essential in supporting them to take on careers in global surgery. These results advocate for the inclusion of pre-clinical global surgery seminars alongside equitable international clinical internships in formal medical education across Europe and worldwide.

## Introduction

Global surgery is “an area of study, research, practice, and advocacy that seeks to improve health outcomes and achieve health equity for all people who require surgical care, with a special emphasis on underserved populations and populations in crisis” [1, 2]. Despite 16.9 million deaths occurring annually due to conditions requiring surgical care, substantially surpassing the number of deaths due to HIV/AIDS, tuberculosis and malaria combined, global surgery can still be regarded as the “neglected stepchild of global health” [3,4].

In 2015, the landmark Lancet Commission on Global Surgery 2030 highlighted important inequalities in access to surgical care worldwide, especially in low- and middle-income countries (LMIC) [2]. Each year, five billion people lack access to safe and affordable surgical and anaesthetic care, 33 million people face catastrophic health expenditure due to out of pocket payments and an additional 48 million due to non-medical costs related to surgical care. Basic surgical, anaesthetic and obstetric care in underserved populations could prevent 1.4 million deaths and 77.2 million Disability Adjusted Life Years (DALYs) annually [5]. Despite this knowledge, many misconceptions remain regarding surgery in LMICs as too expensive, complicated and inefficient compared to other facets of global health [2, 6, 7].

An estimated 2.28 million additional surgeons, anaesthesiologists and obstetricians are needed by 2030 to provide the additional 143 million surgical procedures needed annually [2]. An understanding of how medical students perceive the role of surgery in global health will therefore be essential in developing meaningful educational opportunities for the next generation of the global surgery workforce [7]. Knowledge of the obstacles faced by medical students when pursuing a career in global surgery will also be crucial in providing avenues of career development [8]. The few studies that measured the knowledge, attitudes and exposure of medical students towards global surgery have highlighted a discrepancy between medical students’ interest in global surgery and their lack of knowledge and exposure to this rapidly evolving field [7-11].

The aim of this study is to investigate the knowledge, attitudes and exposure of Belgian medical students towards global surgery and identify possible approaches to global surgery curricula development.

## Material and methods

This study was approved by the institutional review board of the Ghent University Hospital (BC-10194). The survey was developed following a literature review to collect global surgery statistics and identify relevant questionnaires [1, 2, 7-9, 12]. The draft survey was reviewed by three attending surgeons, five medical students and members of the International Student Surgical Network (InciSion) Belgium, a student- and resident-led non-profit organisation regarding global surgery [13, 14]. The final survey created in Qualtrics software © 2022 (Provo, Utah, USA) provided a definition of global surgery and consisted of 28 main and 17 additional questions on demographics, exposure, interest, knowledge, obstacles and attitudes towards global surgery [1, 2]. Questions had multiple choice, free text, checklist, yes or no, ranking (one being the highest and seven the lowest) and Likert scale answers (S1 Appendix).

The anonymous online survey was distributed to first to final year medical students across seven Belgian universities using social media. All participants provided written informed consent for anonymous data collection, storage and analysis. All data was handled confidentially and evaluated by researchers for scientific purposes only.

Data was analysed using RStudio statistics version 2022.02.3 (R Foundation, Vienna, Austria). Descriptive statistics and crude odds ratios (OR) with 95% confidence interval (CI) analysis between stage of medical education, gender and 12 selected survey questions were estimated using the “oddsratio.wald” function in RStudio. Means and standard deviations (SD) for rankings were calculated using the “sd” function in RStudio. Data from medical schools with less than 10 study participants were excluded from the analysis to maintain a sufficient sample size per institution [15].

## Results

### Demographics

A total of 304 responses were included from four universities: Ghent University (57.2%, n = 174/304), the University of Antwerp (20.7%, n = 63/304), the Catholic University of Leuven (12.2%, n = 37/304) and the University of Liège (9.9%, n = 30/304). Participants were first (14.5%), second (16.8%), third (24.7%), fourth (15.8%), fifth (17.1%) and final year (11.2%) medical students. There were more female than male participants (64.8% vs 34.2%). Odds ratios with 95% CI analysis between stage of medical education (bachelor’s/master’s), gender (male/female) and 12 questions on attitudes, exposure and knowledge on global surgery are presented in Table 1.

**Table 1.**
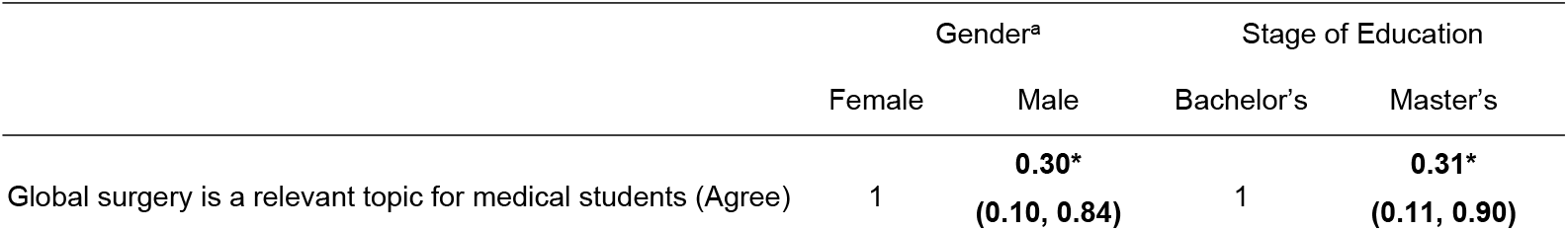
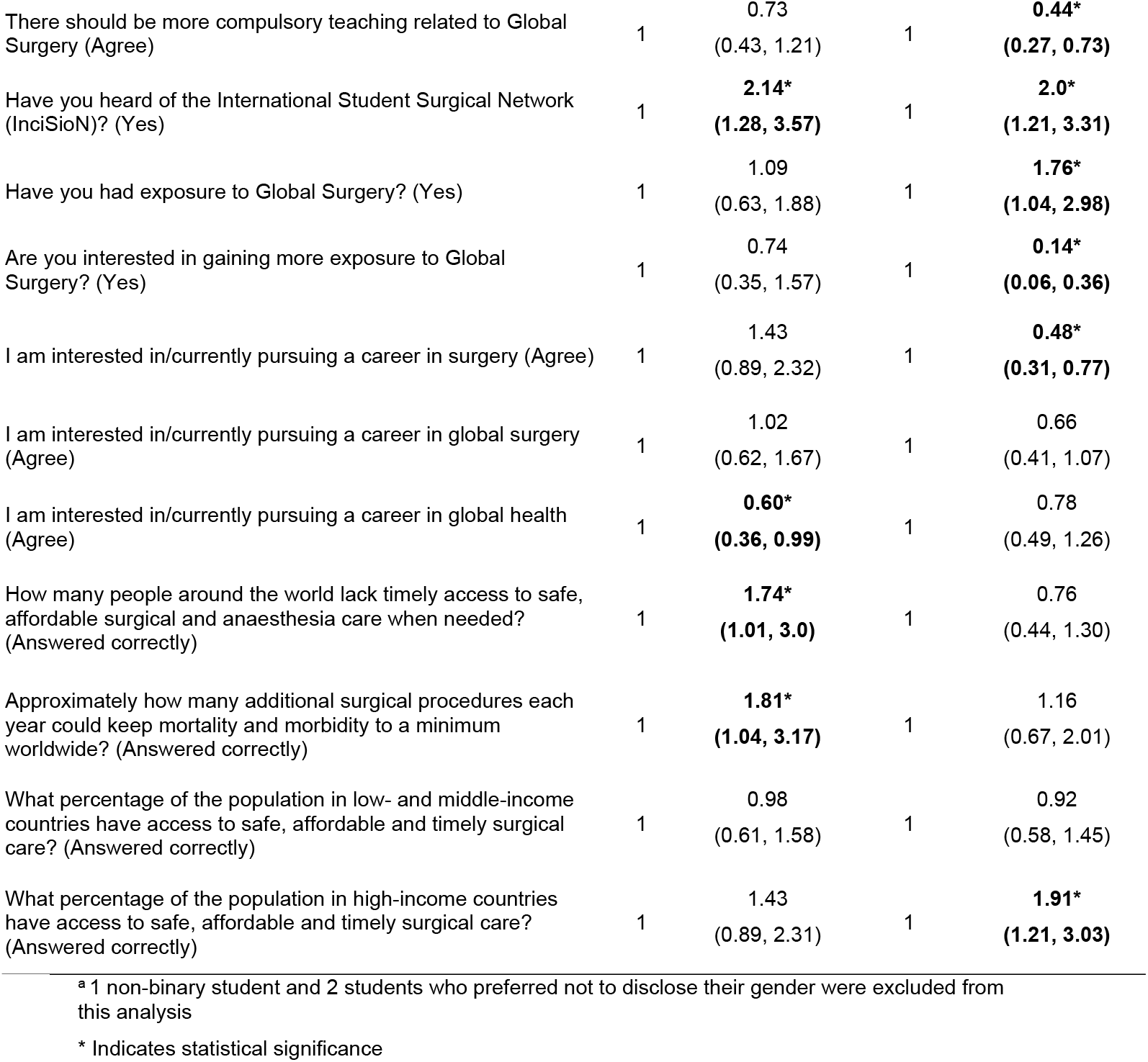
Association between (OR and 95% CI) gender and stage of education and attitudes, exposure and knowledge on global surgery

### Exposure and interest

A minority of students reported having been exposed to global surgery (24.7%, n = 75/304), with social media being the most frequent medium among those exposed (62.7%, n = 47/75) (Figure 1). Master’s students were almost twice as likely to have had exposure than bachelor’s students (OR 1.76). Few students had heard of InciSioN (28.6%, n = 87/304) and even less had taken part in any of InciSioN’s activities (2.3%, n = 7/304). Male students and master’s students were twice likely to have heard of InciSioN (OR 2.14 and 2.0, respectively). Most students were interested in gaining more exposure to global surgery (75.3%, n = 223/296) but master’s students were less likely to be interested (OR 0.14). The most preferred methods of additional exposure were optional teaching programmes at medical school (55.7%, n = 165), events (53%, n = 157), clinical internships (51%, n = 151) and required teaching programmes (45.3%, n = 134) (Figure 1). When asked to rank which of seven global surgery aspects they would like to learn more about, students scored clinical aspects the highest (1.5 ± 1.08) followed by educational (3.72 ± 1.76), cultural (3.78 ± 1.63), ethical (4.11 ± 1.65), organisational (4.71 ± 1.61), research (4.95 ± 1.79) and economic aspects (5.23 ± 1.8).

**Fig. 1.**
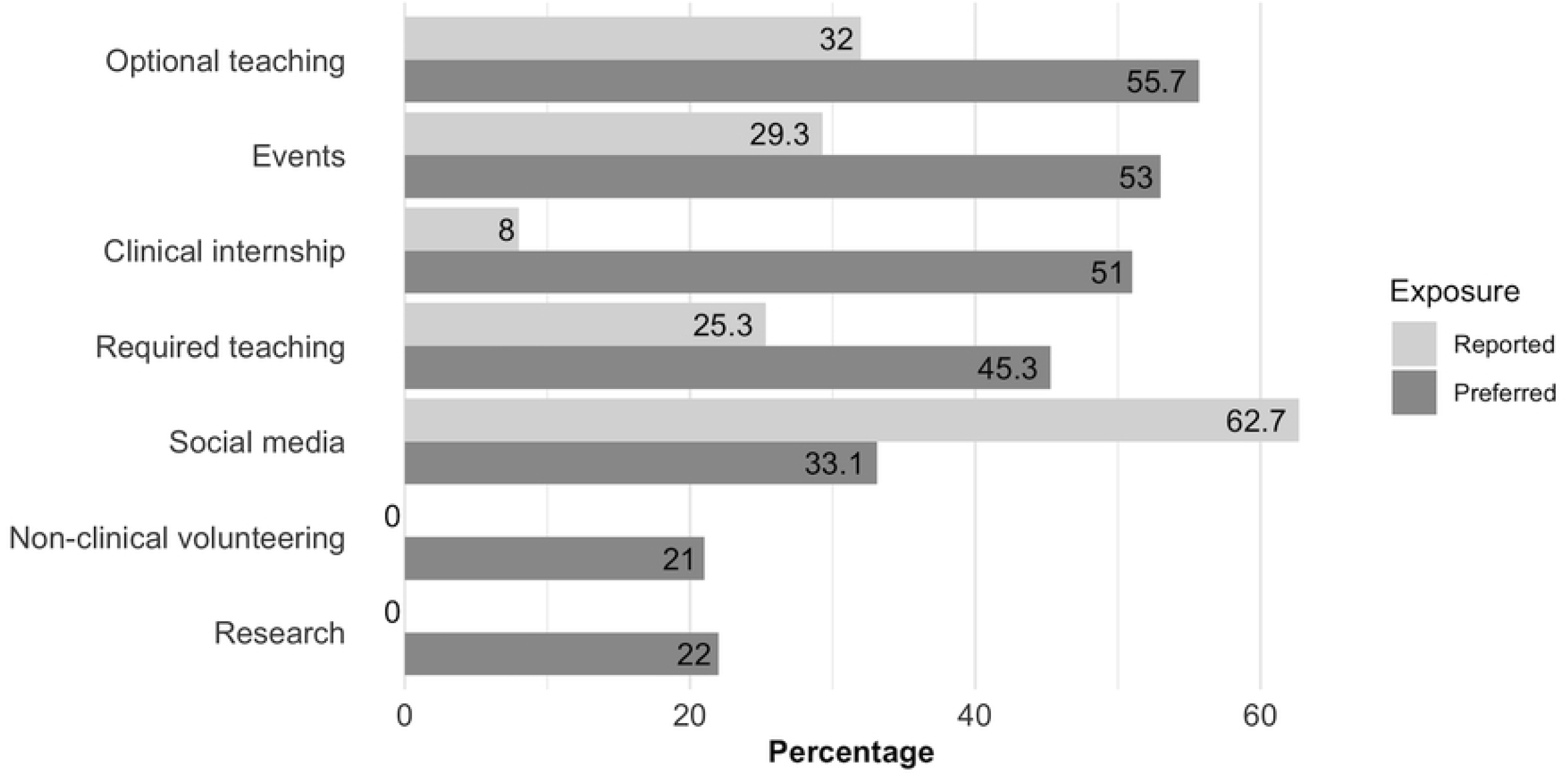
Reported versus preferred methods of global surgery exposure

### Attitudes and education

More students were moderately (43.1%, n = 131/304) or slightly familiar (39.8%, n = 121/304) with global health while only slightly familiar (37.8%, n = 115/304) or not familiar at all (52%, n = 158/304) with global surgery. Most students reported that global health was sometimes (51%, n = 155/304) or rarely (35.5%, n = 108/304) incorporated while global surgery was only rarely (57.9%, n = 176/304) or never incorporated (24%, n = 73/304) into their required medical school curriculum. While almost all students agreed [strongly (49.3%, n = 150/304) or somewhat (45%, n = 137/304)] that global surgery is a relevant topic for medical students, male students compared to female and master’s compared to bachelor’s students were less likely to agree (OR 0.30 and 0.31 respectively). Most students agreed [strongly (26.6%, n = 84/304) or somewhat (43.4%, n = 132/304)] that there should be more compulsory teaching related to global surgery (Figure 2), but master’s students were less interested in a compulsory global surgery curriculum (OR 0.44).

**Fig. 2.**
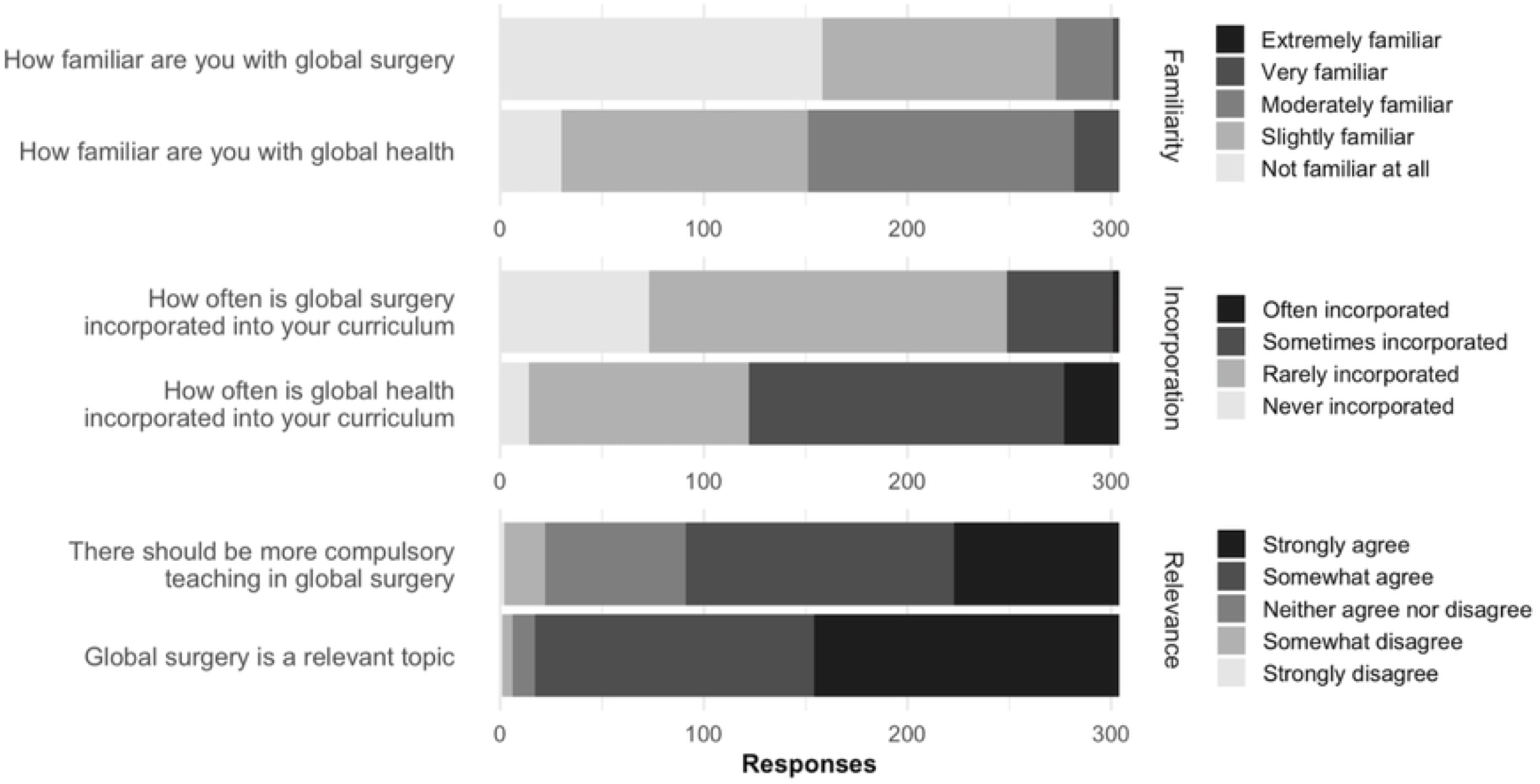
Familiarity, relevance and incorporation of global surgery in student’s medical education

### Knowledge

Overall, knowledge on global surgery was poor. Only 23.9% of students (n = 72/304) could correctly identify that five billion people lack access to timely, safe and affordable surgical care and only 21.7% (n = 66/304) knew that approximately 150 million additional surgical procedures are needed annually. Male students were more likely to answer these two questions correctly (OR 1.74 and 1.81). Less than half of students (44.4%, n = 135/304) correctly identified that only 10 to 30% of the population in LMICs have access to timely, safe and affordable surgical care and only 44.1% of students (n = 134/304) were aware that over 80% of the population in high-income countries (HIC) have access to surgical care. Few (12.8%, n = 39/304) knew the three Bellwether procedures and only a minority of students (21%, n = 64/304) could identify main stakeholders in global surgery (Table 2).

**Table 2.** Responses to knowledge-based global surgery questions^a^

|  | Correct | Incorrect | "I do not know" |
| --- | --- | --- | --- |
| Approximately how many people around the world lack timely access to safe, affordable surgical and anaesthesia care when needed? | 24% | 51% | 25% |
| Approximately how many additional surgical procedures each year could keep mortality and morbidity to a minimum worldwide? | 22% | 23% | 45% |
| What percentage of the population in low- and middle-income countries have access to safe, affordable and timely surgical care? | 44% | 33% | 22% |
| What percentage of the population in high-income countries have access to safe, affordable and timely surgical care? | 44% | 40% | 15% |
| Which of these is NOT a Bellwether procedure? | 13% | 4% | 84% |
| Which of the following is NOT a main stakeholder in global surgery? | 21% | 11% | 68% |

<sup>a</sup> Responses rounded to the nearest number

### Aspirations and obstacles

About half of the students were interested in pursuing a career in surgery (53.3%, n = 162/304), though master’s students were less likely to than bachelor’s students (OR 0.48). More than one in three were interested in a career in global surgery (35.9%, n = 109/304) or global health (37.5%, n = 114/304). Male students were less likely to pursue global health as a career (OR 0.60). Students were not (57.2%, n = 174/304) or only slightly familiar (29.9%, n = 91) with what global surgery means as a career, and most students found pursuing a career in global surgery to be only slightly (45.7%, n = 139/304) or moderately feasible (44.7%, n = 136/304). The most commonly perceived obstacles to a career in global surgery were personal or family responsibilities (42.8%, n = 130/304), language barriers (42.1%, n = 128/304), lack of established career paths in global surgery (39.1%, n = 119/304), financial constraints (34.5%, n = 105/304), and a lack of surgical role models and mentorship (33.6%, n = 102/304) (Figure 3).

**Fig. 3.**
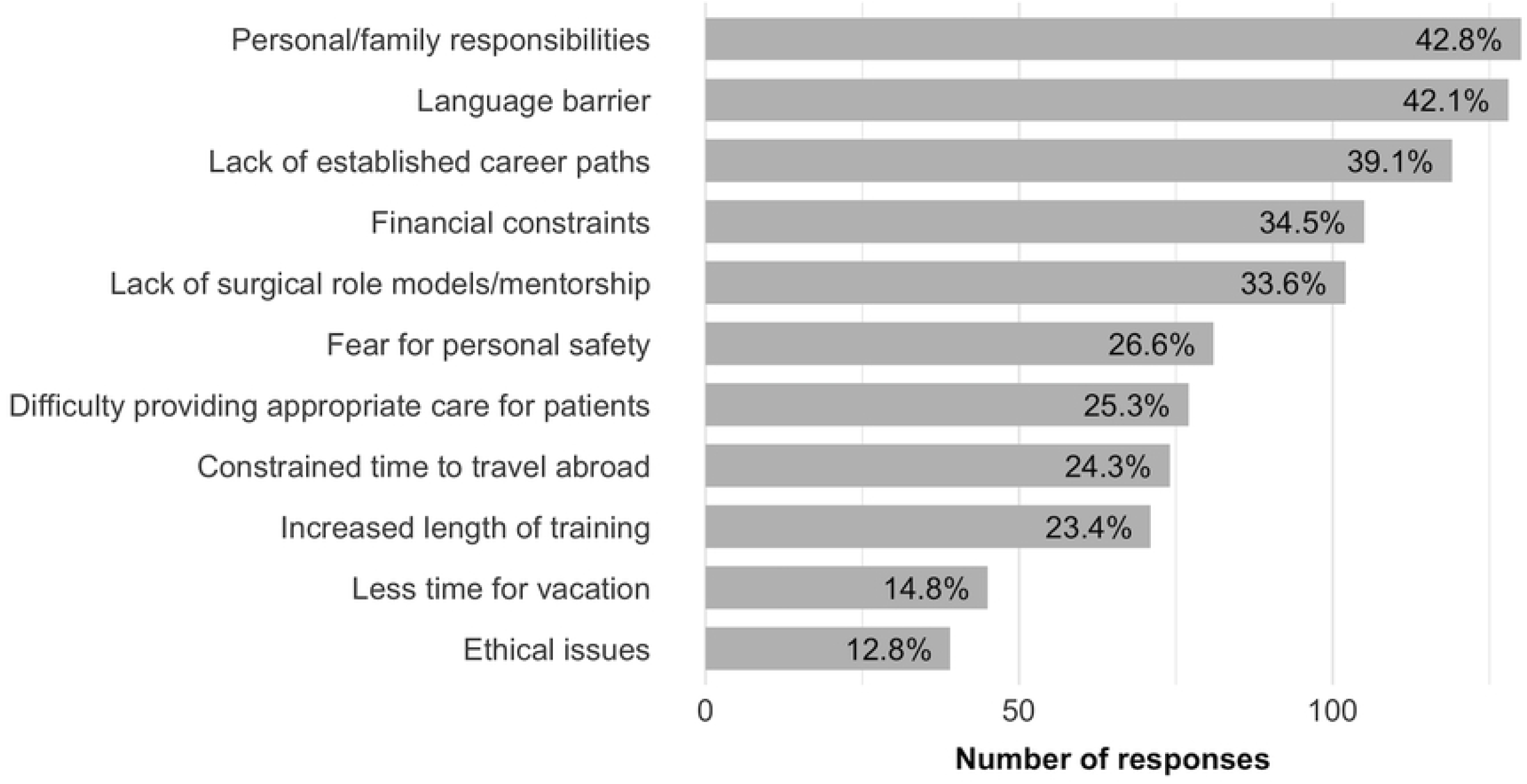
Reported obstacles to pursuing a career in global surgery

## Discussion

To our knowledge this is the first study to quantitatively assess the current knowledge, attitudes and exposure of medical students towards global surgery in a European setting. We found basic knowledge regarding global surgery to be lacking despite a profound interest among students. Despite almost all medical students finding it to be a relevant topic and many wanting more global surgery education, few had been exposed to global surgery. Multiple barriers to pursuing a career in global surgery were reported.

Grasping a basic knowledge of global surgery as an essential component of global health is a first step for medical students in recognizing the urgent need for safe and affordable surgical care worldwide [2]. Belgian students were less familiar with global surgery than with global health and had poor knowledge on its key aspects. Our findings of a lack of knowledge among students are similar to those observed in the UK, USA and LMICs, but results are challenging to compare due to a lack of standardized questions [7-11]. This lack of knowledge likely feeds the misconceptions of global surgery being too specific, not tackling a significant burden of disease or not being cost-effective [7, 10, 16]. Providing all medical students with only a rudimental understanding of the inequalities in surgical care could already be a catalyst towards expanding the global surgery community [7, 10, 17]. This lack of knowledge was echoed by a general lack of exposure with only one in four students being exposed. Despite students’ interest, global surgery exposure opportunities for medical students currently remain limited [17]. Allowing diverse and accessible global surgery exposure from early on in a medical student’s career is critical to increasing their participation as residents and doctors [11, 17].

Involving and educating medical students will form a crucial component in increasing the global surgery workforce by 2.28 million by 2030, one of the goals set by the Lancet Commission on Global Surgery [2, 17]. Most (95%) students agreed that global surgery is a relevant topic and 70% agreed that there should be more compulsory teaching on this subject. Three-quarters of students were interested in gaining more exposure to global surgery. Despite social media being the most common method of previous exposure, students did not prefer this method. They preferred optional teaching, events, clinical internships and required teaching as possible exposure opportunities. This reinforces the notion that students are a requesting party in integrating global surgery as a component of formal medical education [11, 18]. Nevertheless, global surgery was rarely incorporated into Belgian medical curricula, placing a burden on local stakeholders and initiatives to create global surgery workshops, lectures and conferences. Our findings reflect those in other countries, where medical schools have historically struggled to incorporate global surgery and global health into their curricula [11, 17, 19]. This increased understanding of students’ desires will be fundamental in guiding the development of meaningful exposure opportunities in the future [17, 20].

Few established pre-clinical courses on global surgery exist [21, 22]. Novel global surgery curricula have emphasized a tailored approach based on a student’s progression through medical school [19, 21-24]. In our study, master’s students were less interested in additional exposure to global surgery, less likely to agree that it is a relevant topic and less likely to want compulsory global surgery education. This confirms that seminars providing basic knowledge, tackling common misconceptions and discussing cost-effectiveness, ethical considerations and health systems management should be primarily directed towards bachelor’s students [7, 19]. These pre-clinical seminars could be complemented by international internship opportunities to motivated students in the clinical years [7, 17, 23]. Current international internship opportunities are often short and unilateral, running the risk of straining local health systems and harming patients or falling into medical tourism and neo-colonialism [19, 21, 24]. Instead, clinical global surgery internships should be equitable, bidirectional and beneficial for both visitors and hosts [24-27]. To achieve this, providing appropriate pre-departure training and guidance in collaboration with the host country should be an ethical imperative [7, 24].

Students reported several obstacles to pursuing a career in global surgery, primarily personal responsibilities, language barrier, lack of established career paths, financial constraints and a lack of role models/mentorship. These findings are dissimilar to those reported by Mehta et al., who found common obstacles to be constrained time to travel, increased length of training, providing care in resource-limited settings and a lack of established career paths, respectively [7]. While personal responsibilities are inevitable, preparatory language courses and financial support could be provided. The recommendations of the Global Surgery 2030 roadmap for HIC schools and training programs are especially relevant, advocating to “endorse global surgery as an established career path for trainees” and “provide logistical, financial and mentorship support for trainees interested in pursuing global surgery endeavours” [28]. Creating clear career paths and opportunities while providing a broad support and mentorship network will be crucial in lowering the threshold for medical students to aspire for careers within global surgery.

We acknowledge limitations to this study. The population was not a random sample of all medical students enrolled at Belgian universities. Only four of the seven contacted institutions were included, three of the four being in Flemish speaking regions, and not all medical students at each university responded. This possibly introduced a selection bias with students with greater interest being more likely to participate. The proportion of students having exposure and wanting additional exposure may therefore be an overestimate. In addition, there is no validated questionnaire to assess knowledge, attitudes and exposure to global surgery. As such our results are challenging to compare to previous literature. Lastly, results are not directly generalizable to other countries, especially countries outside of Europe.

## Conclusions

In conclusion, Belgian medical students’ knowledge and exposure towards global surgery is lacking despite a profound interest in the field. Medical students across the globe will be tomorrow’s 2.28 million physicians needed in the global surgery workforce of 2030, providing increased access to safe, timely and affordable surgical, anaesthetic and obstetric care for all. Local, national and international global surgery curricula development will be essential to support these students in taking on careers in global surgery. These results advocate for the inclusion of pre-clinical global surgery seminars alongside equitable and bilateral international clinical internships in formal undergraduate medical education in Europe and worldwide.

## Data Availability

The data file and code used for the statistical analysis can be found at: DOI: 10.17632/93gwsz8xt4.2

https://data.mendeley.com/datasets/93gwsz8xt4/2

## Acknowledgements

We would like to thank the InciSioN Belgium team and the Belgian Medical Student’s Association for their assistance in disseminating the survey.

## Supporting information caption

S1 Appendix: Online survey for medical students regarding global surgery

## Notes

### Competing Interest Statement

The authors have declared no competing interest.

### Funding Statement

The authors received no specific funding for this work.

### Author Declarations

This study was approved by the institutional review board of the Ghent University Hospital (BC-10194).

## References

1. Dare AJ, Grimes CE, Gillies R, Greenberg SL, Hagander L, Meara JG, et al. Global surgery: defining an emerging global health field. Lancet. 2014;384:2245–2247. DOI: 10.1016/S0140-6736(14)60237-3

2. Meara JG, Greenberg SL. The Lancet Commission on Global Surgery Global surgery 2030: Evidence and solutions for achieving health, welfare and economic development. Surgery. 2015;157:834–835. DOI: 10.1016/j.surg.2015.02.009

3. Farmer PE, Kim JY. Surgery and global health: a view from beyond the OR. World J Surg. 2008;32:533–536. DOI: 10.1007/s00268-008-9525-9

4. Lozano R, Naghavi M, Foreman K, Lim S, Shibuya K, Aboyans V, et al. Global and regional mortality from 235 causes of death for 20 age groups in 1990 and 2010: a systematic analysis for the Global Burden of Disease Study 2010. Lancet. 2012;380:2095–2128. DOI: 10.1016/S0140-6736(12)61728-0

5. Bickler SN, Weiser TG, Kassebaum N, Higashi H, Chang DC, Barendregt JJ, et al. Global Burden of Surgical Conditions. In: Debas HT, Donkor P, Gawande A, Jamison DT, Kruk ME, Mock CN, eds. Essential Surgery: Disease Control Priorities, Third Edition (Volume 1). Washington (DC): The International Bank for Reconstruction and Development / The World Bank © 2015 International Bank for Reconstruction and Development / The World Bank.; 2015.

6. Farmer P. Dispelling myths about surgery’s role in global health. Investing in Health: World Bank Blogs; 2015. Accessed 9 October 2022. https://blogs.worldbank.org/health/dispelling-myths-about-surgery-s-role-global-health.

7. Mehta A, Xu T, Murray M, Casey KM. Medical Student Perceptions of Global Surgery at an Academic Institution: Identifying Gaps in Global Health Education. Acad Med. 2017;92:1749–1756. DOI: 10.1097/ACM.0000000000001832

8. International Survey of Medical Students Exposure to Relevant Global Surgery (ISOMERS): A Cross-Sectional Study. World J Surg. 2022;46:1577–1584. DOI: 10.1007/s00268-022-06440-0

9. Kanmounye US, Mbonda AN, Djiofack D, Daya L, Pokam OF, Ghomsi NC. Exploring the knowledge and attitudes of Cameroonian medical students towards global surgery: A web-based survey. PLoS One. 2020;15:e0232320. DOI: 10.1371/journal.pone.0232320

10. Abraham MN, Abraham PJ, Chen H, Hendershot KM. What is global surgery? Identifying misconceptions among health professionals. Am J Surg. 2020;220:271–273. DOI: 10.1016/j.amjsurg.2019.11.021

11. Scott EM, Fallah PN, Blitzer DN, NeMoyer RE, Sifri Z; Global Surgery Student Alliance (GSSA), et al. Next Generation of Global Surgeons: Aligning Interest With Early Access to Global Surgery Education. J Surg Res. 2019;240:219–226. DOI: 10.1016/j.jss.2019.03.009

12. Alkire BC, Raykar NP, Shrime MG, Weiser TG, Bickler SW, Rose JA, et al. Global access to surgical care: a modelling study. Lancet Glob Health. 2015;3:e316–323. DOI: 10.1016/S2214-109X(15)70115-4

13. International Student Surgical Network (InciSioN). 2022. Accessed 9 October 2022. https://incisionetwork.org/.

14. Vervoort D, Bentounsi Z. InciSioN: Developing the Future Generation of Global Surgeons. J Surg Educ. 2019;76:1030–1033. DOI: 10.1016/j.jsurg.2019.02.008

15. Rennie N, Degraeuwe E, Deltour C, et al. Students’ perspectives on global surgery and its integration into formal medical education. Mendeley Data. Version 2. 21 November 2022. DOI: 10.17632/93gwsz8xt4.2

16. Bae JY, Groen RS, Kushner AL. Surgery as a public health intervention: common misconceptions versus the truth. Bull World Health Organ. 2011;89:394. DOI: 10.2471/BLT.11.088229

17. Gosselin-Tardif A, Butler-Laporte G, Vassiliou M, Khwaja K, Ntakiyiruta G, Kyamanywa P, et al. Enhancing medical students’ education and careers in global surgery. Can J Surg. 2014;57:224–225. DOI: 10.1503/cjs.027713

18. Patel R, Khundkar R, Peter N, Turner J, Edgcombe H, Makins A, et al. Improving global surgery education for trainees. IJS Global Health. 2019;2. DOI: 10.1097/GH9.0000000000000007

19. Moren A, Cook M, McClain M, Doberne J, Kiraly L, Perkins RS, et al. A pilot curriculum in international surgery for medical students. J Surg Educ. 2015;72:e9–e14. DOI: 10.1016/j.jsurg.2015.04.027

20. Tannan SC, Gampper TJ. Resident Participation in International Surgical Missions is Predictive of Future Volunteerism in Practice. Arch Plast Surg. 2015;42:159–163. DOI: 10.5999/aps.2015.42.2.159

21. Anderson GA, Albutt K, Holmer H, Muguti G, Mbuwayesango B, Muchuweti D, et al. Development of a Novel Global Surgery Course for Medical Schools. J Surg Educ. 2019;76:469–479. DOI: 10.1016/j.jsurg.2018.07.026

22. Jayaram A, Pawlak N, Kahanu A, Fallah P, Chung H, Valencia-Rojas N, et al. Academic Global Surgery Curricula: Current Status and a Call for a More Equitable Approach. J Surg Res. 2021 Nov;267:732–744. doi: 10.1016/j.jss.2021.03.061. DOI: 10.1016/j.jss.2021.03.061

23. Kühner S, Ekblad S, Larsson J, Löfgren J. Global surgery for medical students - is it meaningful? A mixed-method study. PLoS One. 2021;16:e0257297. DOI: 10.1371/journal.pone.0257297

24. Zivanov CN, Joseph J, Pereira DE, MacLeod JBA, Kauffmann RM. Qualitative Analysis of the Host-Perceived Impact of Unidirectional Global Surgery Training in Kijabe, Kenya: Benefits, Challenges, and a Desire for Bidirectional Exchange. World J Surg. 2022;46:2570–2584. DOI: 10.1007/s00268-022-06692-w

25. Grant CL, Robinson T, Al Hinai A, Mack C, Guilfoyle R, Saleh A. Ethical considerations in global surgery: a scoping review. BMJ Glob Health. 2020;5:e002319. DOI: 10.1136/bmjgh-2020-002319

26. Bozinoff N, Dorman KP, Kerr D, Roebbelen E, Rogers E, Hunter A, et al. Toward reciprocity: host supervisor perspectives on international medical electives. Med Educ. 2014;48:397–404. DOI: 10.1111/medu.12386

27. Kumwenda B, Dowell J, Daniels K, Merrylees N. Medical electives in sub-Saharan Africa: a host perspective. Med Educ. 2015;49:623–633. DOI: 10.1111/medu.12727

28. Ng-Kamstra JS, Greenberg SLM, Abdullah F, Amado V, Anderson GA, Cossa M, et al. Global Surgery 2030: a roadmap for high income country actors. BMJ Glob Health. 2016;1:e000011. DOI: 10.1136/bmjgh-2015-000011

